# Normal feeding movements expressed by dimensionality reduction of whole-body joint motions using principal component analysis

**DOI:** 10.1101/2024.09.14.24313686

**Authors:** Jun Nakatake, Shigeaki Miyazaki, Hideki Arakawa, Etsuo Chosa

## Abstract

**Background:** Understanding elementary feeding movements and postures is essential for improving assessment and intervention strategies in occupational therapy, particularly for individuals with eating difficulties, and for educating caregivers and students. However, the current assessment tools lack precision in evaluating complex feeding movements and often rely on subjective judgments rather than objective measures. We aimed to determine elementary movements and postures corresponding to different feeding phases using principal component analysis (PCA).

**Methods:** This cross-sectional observational study was conducted at a Local National University Hospital and included 45 healthy, right-handed adult volunteers (23 men and 22 women) aged 20–39 years (mean age, 27.3 years), with no neurological or musculoskeletal impairments. Movements during yogurt feeding using a spoon were captured with a three-dimensional inertial sensor motion capture system. Principal components (PCs) and their scores were derived from PCA of whole-body joint motion data across four feeding phases. PC scores were compared between phases using Friedman’s and post-hoc tests.

**Results:** The primary PC, representing whole-body movement, accounted for 50.0% of the variance; the second PC, associated with hand direction changes, accounted for 13.7%. The cumulative variance of the first six PCs was 87.4%, including individual body-part movements and fixations or combinations of these. Significant differences existed between feeding phases, particularly in the reaching and transport phases, which showed greater whole-body movement than that during the spooning and mouth phases. Hand direction changes were more prominent during the spooning phase than during the mouth phase.

**Conclusions:** PCA helped determine key elementary movements and their corresponding feeding phases, which can be used to assess patients with feeding difficulties and guide occupational therapy interventions.

## Introduction

Eating difficulties caused by injuries or illnesses result in physiological, psychological, and social challenges (Cipriano-Crespo et al., 2020; Klinke et al. 2013). Impaired self-feeding skills are further associated with malnutrition (Ciliz et al., 2023). Occupational therapy addresses these issues by focusing on swallowing, posture, movement, equipment, care methods, and habits, and providing psychosocial interventions for patients (Boop et al., 2017; Philipps et al. 2012). Several interventions have been developed to improve posture in children (Bhattacharjya et al., 2021; Mlinda, Leyna, & Massawe, 2018) and provide intensive training for specific movements (Jo, Noh, & Kam, 2020; Treger et al., 2012). Additionally, early rehabilitation using feeding devices in intensive care units has been introduced (Koester et al., 2018).

Eating evaluations are often conducted using assessment tools such as the functional independence measure (Uniform Data System for Medical Rehabilitation, 1990) and the modified Barthel index (Shah, Vanclay, & Cooper, 1989), which include eating as one of several daily tasks. Patients are assessed on an ordinal scale based on judgments regarding voluntary movements, caregiver support, or types of equipment used. An eating-specific screening tool, the minimal eating observation form II, has also been developed (Westergren et al., 2009). This tool consists of three items observed on a nominal or ordinal scale, focusing on the sitting position. Practitioners use these assessments to evaluate the eating conditions of patients, set treatment goals, and guide feeding movements and postures. However, targeted movements and postures may be assessed subjectively owing to the experience of practitioners, and the rationale for these assessments remains insufficient.

While studies had reported summarized measurements and waveforms regarding joint angles necessary for normal feeding activities (Nagao, 2004; Doğan et al., 2019; van Andel et al., 2008), these movement and posture patterns correspond to different time phases (Nakatake et al., 2021). The results suggest substantial changes in whole-body joint angles during the phases of reaching for the dish and transporting food to the mouth. Additionally, motion direction varies during the phases of spooning food and taking it into the mouth. Understanding these joint motions, which involves changes in each joint angle over time, can enhance assessments and interventions aimed at improving the movements or postures of patients (Kontaxis et al., 2009). However, relying solely on individual joint motion attached to corresponding time phases to understand feeding movements is insufficient. Practitioners often recognize the movements of patients based on approximate body part motions rather than individual joint motions, which may involve a combination of joint motions, namely coordination. These movements during feeding, such as reaching for food, manipulating the direction of the palm, approaching the food with the mouth, or stabilizing the trunk to support the upper limbs, may involve coordinated, complex joint motions that play significant roles in daily living. However, their specific functions remain unclear.

To address this issue, we employed principal component analysis (PCA) with quantitative kinematic measurements. This method highlights specific fictitious features of data through dimensional reduction, summarizing variables, which is advantageous for analyzing rich biomechanical variables (Daffertshofer et al., 2004). PCA has revealed elementary movement (EM) patterns from gross movement during tasks, such as reaching (Kaminski, 2007) and trunk bending (Tricon et al., 2007). Furthermore, bilateral upper-limb movements (Burns et al., 2017) and postures in sign languages (Bigand et al., 2021) have been categorized into patterns. EM is interpreted as a combination of joint motions for movements that interact with the environment or within the body.

We hypothesized that EMs could be defined from combinations of joint motions during feeding movements and that the appearance of EMs would differ between feeding phases. Therefore, we assessed the secondary analysis of a previous dataset (Nakatake et al., 2021) and aimed to determine the EMs involved in the feeding phases of whole-body joint motion in healthy individuals using PCA. The identified normal feeding movements and postures could provide clinical observational assessments or intervention cues for patients with eating difficulties.

## Materials and methods

### Participants

The study adhered to the STROBE guidelines (von Elm et al., 2007) and was presented in preprint server (Nakatake et al., 2024). The sample for this study was included in a previous study (Nakatake et al., 2021), and the study protocol was approved by the Research Ethics Committee of the Faculty of Medicine at the University of Miyazaki (Miyazaki-shi, Japan) (approval number: O-1501). As we could not contact previous study participants, they were informed about the study through the institution’s website and provided with the option to opt out of participation at any time. Consequently, informed consent was indirectly obtained from participants who did not decline to participate in this study. Furthermore, the authors did not have access to information that could identify individual participants of the previous study after data collection. In total, 50 participants were recruited from our institutional staff from April 2013 to October 2017, meeting the following criteria: aged 20–39 years, right-handed, and without neurological or musculoskeletal impairments. Individuals who were left-handed for regular spoon use were excluded.

### Measurement procedures

The measurement procedures performed in the occupational therapy room at the institution and instrument details have been previously described (Nakatake et al., 2021). The feeding movements of participants were recorded using a three-dimensional motion capture system (Xsens MVN system; Xsens Technologies B.V., Netherlands). This system provides kinematic output of a biomechanical whole-body model composed of 17 inertial sensors attached to the participant’s head, sternum, scapulas, pelvis, upper-arms, forearms, hands, upper-legs, lower-legs, and feet, consisting of 23 body segments, including the head, neck, pelvis, four vertebrae, scapulae, upper arms, forearms, hands, upper legs, lower legs, feet, and toes. The neutral (zero) position of the joint angles was defined as the joint angle when standing upright with feet parallel, one foot width apart, upper limbs alongside the body, palms facing forward, and the head oriented forward.

Participants sat on a stool of height 40 cm without a backrest, with a table adjusted at their elbow height that were positioned in front of their trunk at a distance of 10 cm. They were instructed to use their right hand to reach for yogurt in a bowl placed on the table, scoop it with a stainless spoon (17.5 cm in length, 41 g in weight), transport it, and bring it to their mouth at a comfortable pace, repeating the sequence thrice. The aim was to capture voluntary movements; therefore, movements from after the spoon left the mouth to before the second instance, and from after the second instance to before the third instance, were analyzed.

### Sample selection

Given that participants performed their own feeding movements, five individuals displaying the following movements were excluded from the standardization of normal feeding movements: unnecessary upper limb elevation, shaking yogurt off the spoon while transporting it to the mouth, looking away, shaking the head vertically while reaching for the bowl, repeated spooning, or separating yogurt with the spoon. The final sample included 45 participants (23 men and 22 women) with a mean age of 27.3 years (standard deviation [SD] = 5.1) and an average height of 164.8 cm (SD = 8.6).

### Data analysis

Joint angles during a successive feeding cycle, consisting of reaching the hand to the bowl (reaching phase), spooning yogurt (spooning phase), transporting yogurt to the mouth (transport phase), and bringing yogurt to the mouth (mouth phase), were identified by confirming pictures in recorded movies synchronized to the system and extracted from the biomechanical model. Data were collected at the right shoulder (flexion/extension, abduction/adduction, and internal/external rotation), elbow (flexion/extension), forearm (pronation/supination), wrist (palmar/dorsal flexion and radial/ulnar deviation), C7-T1 (flexion/extension and right/left lateral flexion), and hip (flexion/extension) at a frequency of 120 Hz. A typical case is displayed in Fig 1. The change in joint angles in each feeding phase was calculated using maximum and minimum values. Performance times during the phases were also recorded.

**Fig 1.**
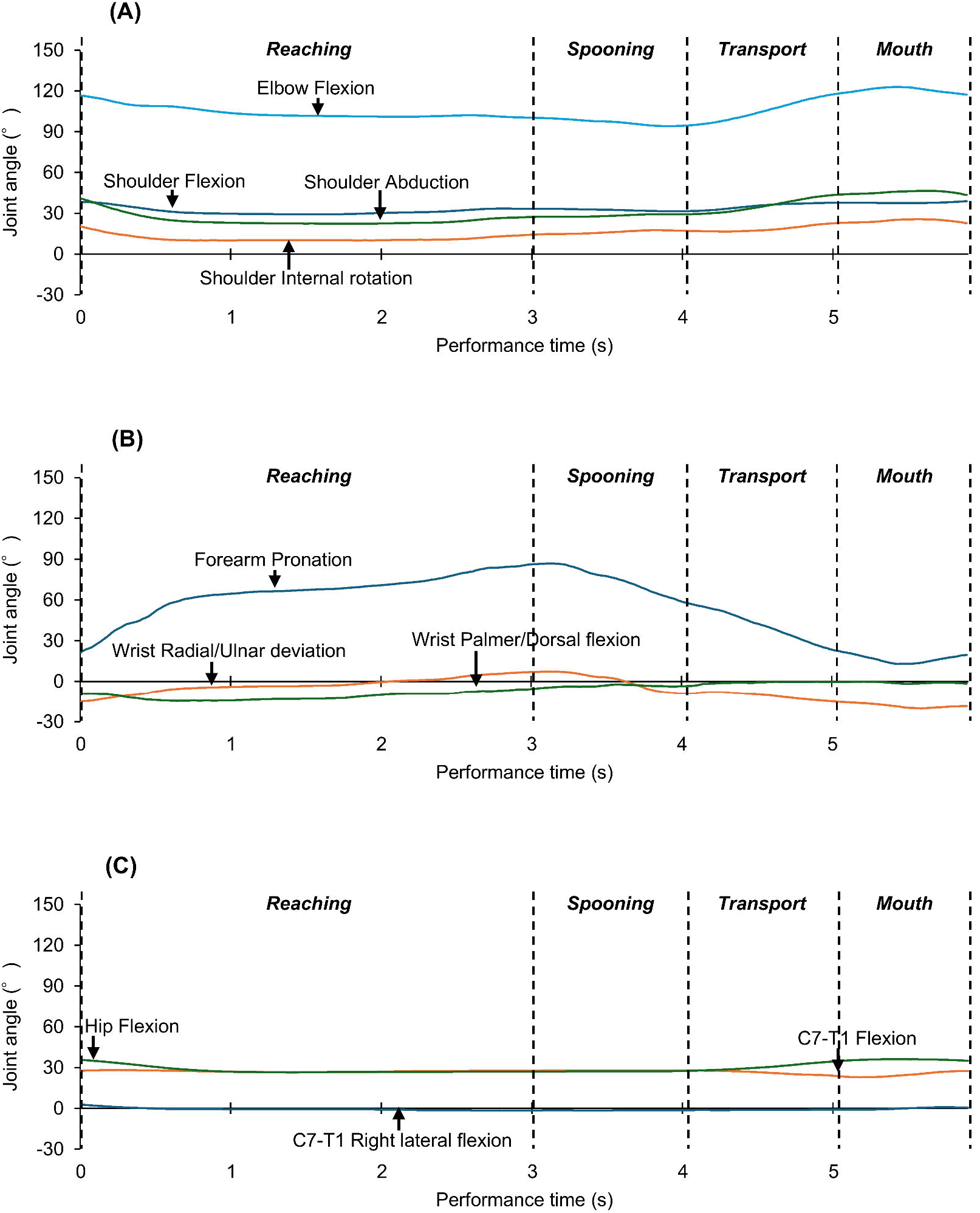
Typical waveforms of whole-body joint angle change across all feeding phases. (A) Changes in the shoulder and elbow joint angles. (B) Changes in the forearm and wrist joint angles. (C) Changes in the neck (C7-T1) and hip joint angle. In all figures, the vertical axis indicates the joint angle, the horizontal axis indicates the performance time, and the vertical dotted line indicates the initial and final durations of each feeding phase.

This study performed PCA which summarizes entire variables to theorical fictitious components called principal components (PCs). PCs consist of sum of substantial measurements loaded by coefficients. Accordingly, each data point has scores of PC. To test our hypothesis, we analyzed changes in joint angles and performance times (11 variables) for one sample containing four feeding phases for each participant (data points of 45 × 4 = 180) using PCA. Subsequently, the PC was selected to ensure that the cumulative variance ratio (contribution ratio to all PC) was ≥ 85%. PCs indicating EMs were interpreted from the loadings. Additionally, PC scores were calculated and compared between feeding phases using the Friedman test, with the significance level set at *p* <.05. Post-hoc analysis was performed using the Wilcoxon signed-rank test with Bonferroni correction (*p* <.008). The effect size *r* was interpreted as medium = |0.3| and large = |0.5| (Hugh & Hugh, 2009).

## Results

### Principal component analysis

The first PC accounted for 50.0% of the variance, followed by the second PC with 13.7%. The variances for PCs 3 through 6 were 7.8%, 6.5%, 5.0%, and 4.3%, respectively. The cumulative variance across the first six PCs totaled 87.4%. The loadings for each PC (Fig 2) allowed for the following interpretations: PC 1 represented whole-body movement over time; PC 2 indicated changes in hand direction while maintaining head stability; PC 3 involved elbow joint motion with stable shoulder joint angles; PC 4 captured lateral neck motion with fixed elbow angles; PC 5 reflected wrist palmar/dorsal flexion; and PC 6 highlighted trunk stability achieved via hip joint fixation.

**Fig 2.**
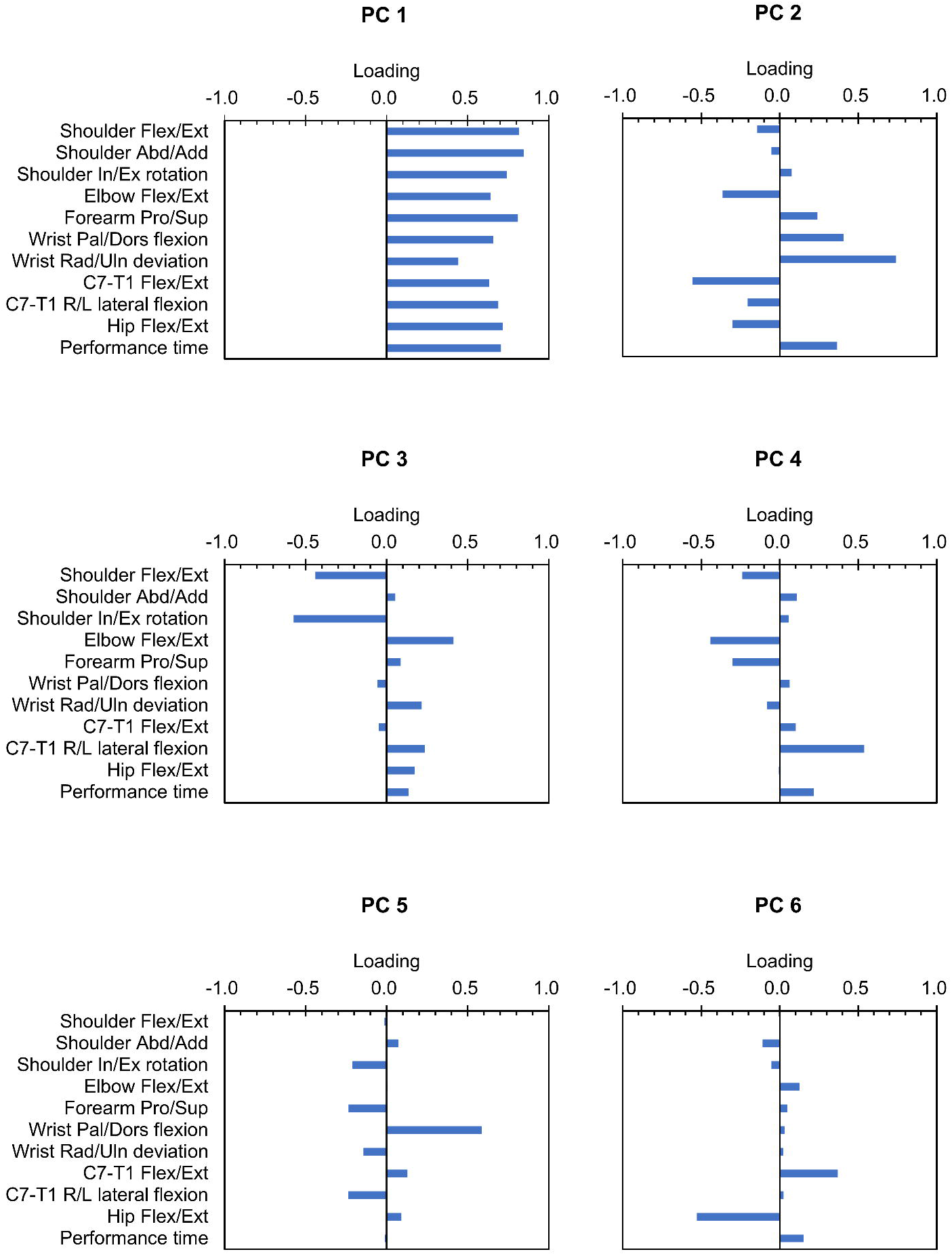
Parameter loadings of PCs. Vertical axis indicates parameters, and horizontal axis indicates loadings. PC, principal component.

### Movements characterizing feeding phases

Friedman’s test revealed significant differences in PC scores across feeding phases for PCs 1–5 (PC 1, χ^2^ = 122, *p* <.0001; PC 2, χ^2^ = 109, *p* <.0001; PC 3, χ^2^ = 32, *p* <.0001; PC 4, χ^2^ = 66, *p* <.0001; PC 5, χ^2^ = 18, *p* =.0004). PC 6 did not show significant differences between phases (χ^2^ = 8, *p* =.0554).

The PC scores for each phase across the first five PCs are summarized in Fig 3 and Tables S2 and S3. PC 1 scores were highest in the reaching phase, followed by the transport, spooning, and mouth phases, with significant differences and large to medium effect sizes between phases (reaching vs. spooning, *z* = -5.8, *p* <.0001, *r* = -0.62; reaching vs. transport, *z* = -5.8, *p* <.0001, *r* = -0.61; reaching vs. mouth, *z* = -5.8, *p* <.0001, *r* = -0.62; spooning vs. transport, *z* = -5.8, *p* <.0001, *r* = -0.62; spooning vs. mouth, *z* = -3.5, *p* =.0005, *r* = -0.36; transport vs. mouth, *z* = -5.8, *p* <.0001, *r* = -0.62).

**Fig 3.**
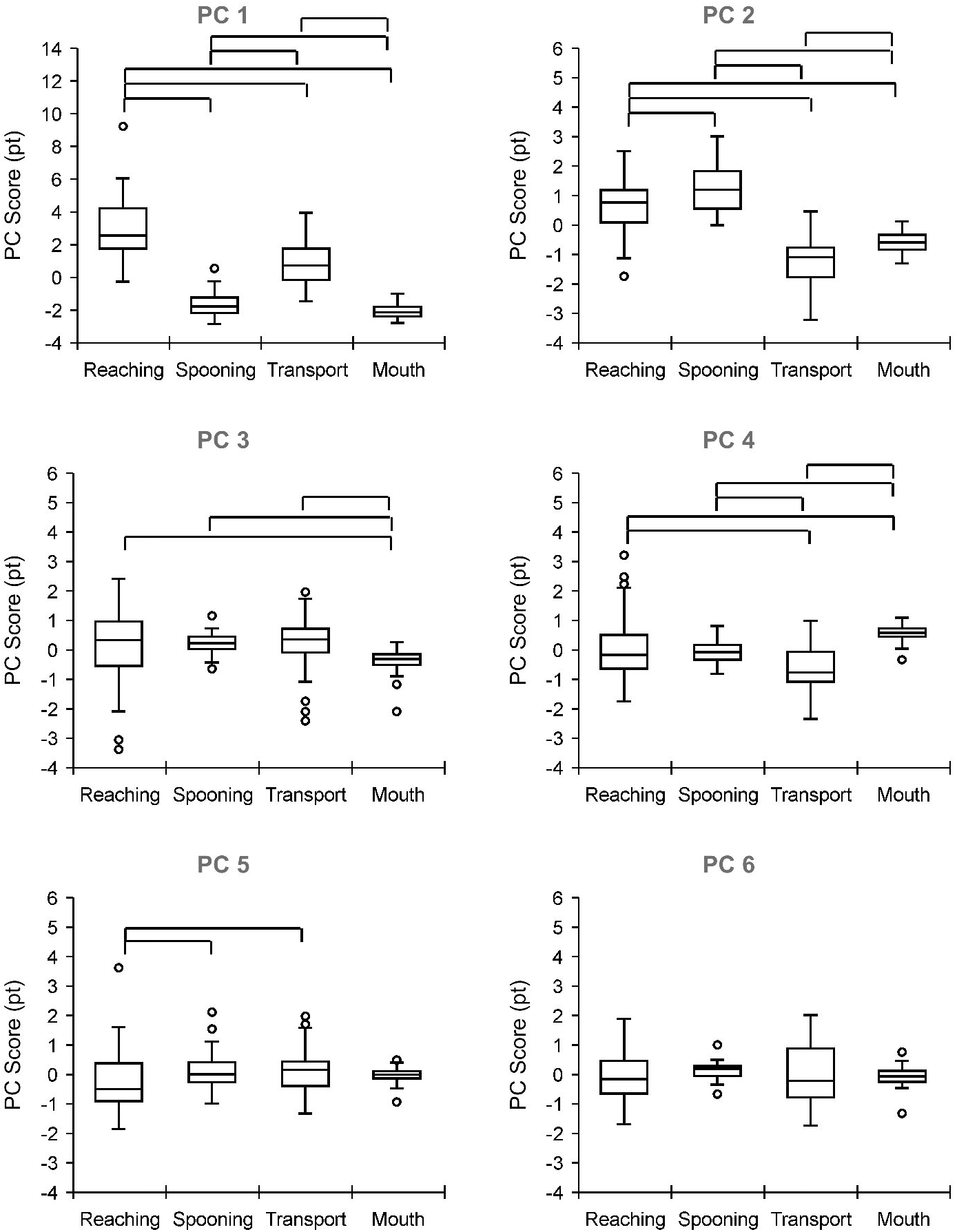
Comparison of PC scores between feeding phases. Vertical and horizontal axes indicate the PC score and feeding phases, respectively. The upper, middle, and lower lines of the boxplot indicate the upper, median, and lower quartiles, respectively. The upper and lower bars indicate the maximum and minimum values, respectively. Dots indicate outliers. Box plots connected by the above lines show significant differences between feeding phases, as determined using post-hoc analysis of the Wilcoxon signed–rank test with Bonferroni correction (*p* <.008). PC, principal component.

For PC 2, scores between all phases were significant with large effect to medium sizes, in the order of spooning, reaching, mouth, and transport phases (reaching vs. spooning, *z* = -3.1, *p* < .0018, *r* = -0.33; reaching vs. transport, *z* = -5.8, *p* <.0001, *r* = -0.62; reaching vs. mouth, *z* = -5.5, *p* <.0001, *r* = -0.58; spooning vs. transport, *z* = -5.8, *p* <.0001, *r* = -0.62; spooning vs. mouth, *z* = -5.8, *p* <.0001, *r* = -0.62; transport vs. mouth, *z* = -4.9, *p* <.0001, *r* = -0.52).

PC 3 scores were higher in the reaching, spooning, and transport phases than in the mouth phase, with large to small effects (reaching vs. mouth, *z* = -2.7, *p* =.0066, *r* = -0.29; spooning vs. mouth, *z* = -5.5, *p* <.0001, *r* = -0.58; transport vs. mouth, *z* = -3.1, *p* =.0019, *r* = -0.33).

For PC 4, the mouth phase had the highest scores (mouth vs. reaching, *z* = -2.8, *p* =.0045, *r* = -0.30; mouth vs. spooning, *z* = -5.6, *p* <.0001, *r* = -0.59; mouth vs. transport, *z* = -5.6, *p* <.0001, *r* = -0.59), whereas the transport phase had the lowest scores (transport vs. reaching, *z* = -5.2, *p* <.0001, *r* = -0.55; transport vs. spooning, *z* = -4.0, *p* =.0001, *r* = -0.43), with significant differences and large or medium effect sizes.

Finally, PC 5 scores were significantly higher in the spooning and transport phases than in the reaching phase, with medium effect sizes (spooning vs. reaching, *z* = -3.3, *p* =.0009, *r* = -0.35; transport vs. reaching, *z* = -3.3, *p* =.0009, *r* = -0.35).

## Discussion

In this study, we aimed to identify elementary feeding movements and postures based on joint kinematics using PCA and to compare these movements across different feeding phases. The analysis revealed that the six PCs accounted for over 85% of the variance across all phases, supporting the hypothesis that EMs are defined by combinations of joint motions during feeding and that their occurrence varies across feeding phases. These findings suggest that understanding EMs can enhance the ability of occupational therapists to assess and improve feeding movements and postures through targeted interventions, such as positioning, specific movement training, and the use of adaptive devices.

The primary EM, involving whole-body movement for mouth and hand coordination, was most prominent in the reaching phase, followed by the transport phase. The second EM, which involved changes in hand direction by coordinating wrist joint motions with the fixed neck flexion angle, was prominent in the spooning phase followed by reaching phase. The third EM, characterized by elbow motion with fixed shoulder angles, was frequently observed in the spooning, transport, and reaching phases. Lateral neck motion with fixed elbow angles was mostly observed in the mouth phase, but not in the transport phase. The spooning and transport phases involved more wrist flexion/extension movements. Trunk stability, achieved through hip joint fixation, was consistently recognized across all phases (Fig 4).

**Fig 4.**
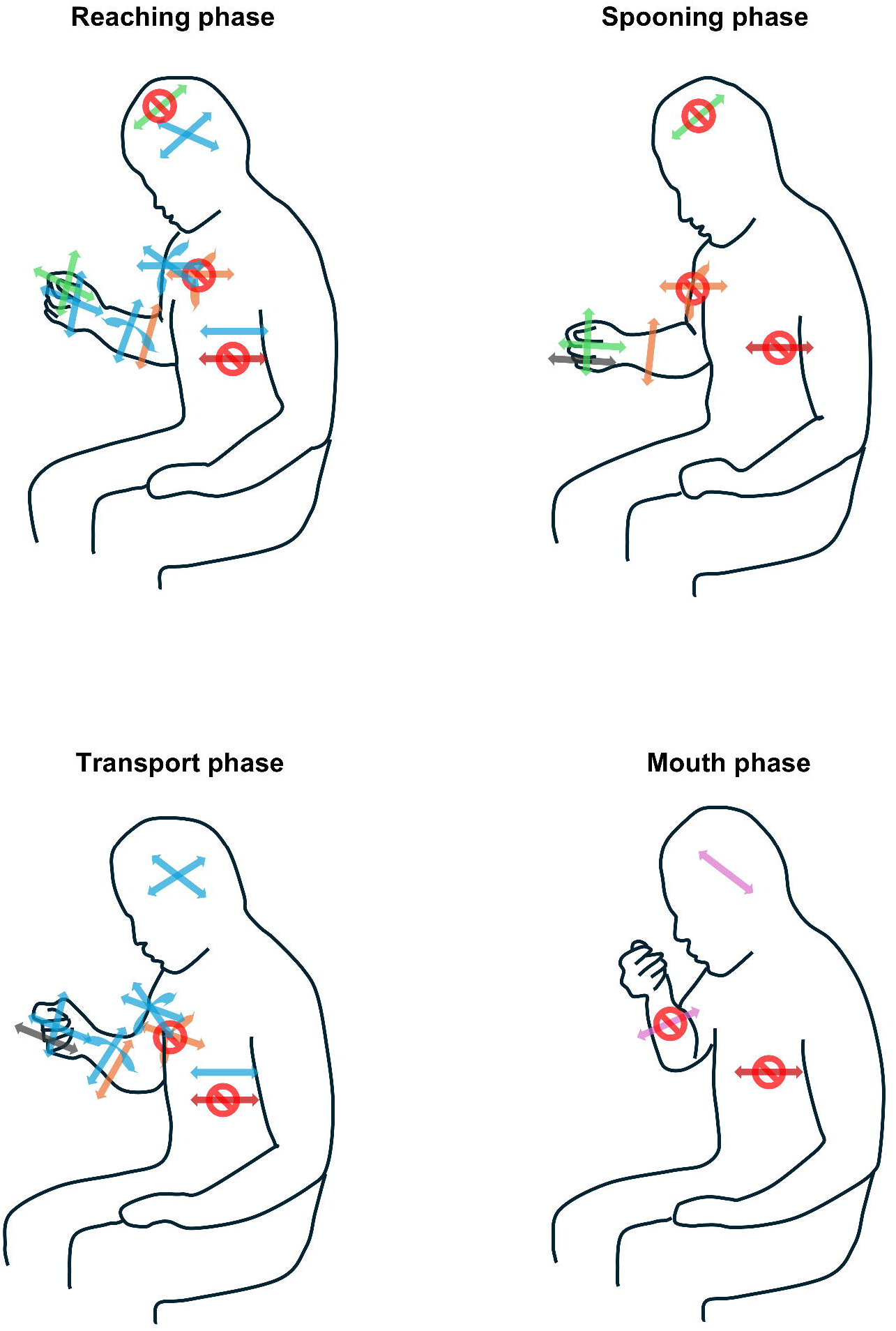
Image of PCs embedded in feeding phases. The line art shows human movements and postures while sitting during each feeding phase. The arrow directions indicate the possibility of motion corresponding to each body part. Red circles with oblique lines indicate that the motion of the overlapping arrows does not occur. Light blue indicates whole-body movements that require time during the reaching and transport phases. Green indicates changing the hand direction while maintaining the neck flexion angle during the reaching and spooning phases. Orange indicates the elbow motion that maintains shoulder flexion and rotation angles during the reaching, spooning, and transporting phases. Pink indicates lateral neck motion while maintaining the elbow angle in the mouth phase. Gray indicates wrist palm/dorsal flexion during spooning, transport, and mouth phases. Brown color indicates trunk posture fixation during all feeding phases. PC, principal component.

The whole-body movement observed in the reaching and transport phases aligns with that reported previously (Nakatake et al., 2021), confirming the coordination of upper and lower limb and neck joint motions. This theorical EM was revealed by the application of PCA to the biomechanical data. During the reaching phase, the shoulder flexes, abducts, and internally rotates, whereas the elbow extends, positioning the hand toward the bowl. The transport phase is characterized by the movement of the upper limb toward the mouth. The range of motion in hip flexion/extension during reaching facilitates the trunk’s return to a neutral position, which is necessary for hand-reaching motion within upper limb length (Kaminski, Bock, & Gentile, 1995). Additionally, head and trunk movements during the transport phase bring the mouth closer to the food (Chinju et al., 2024; Inada et al., 2012; van der Kamp & Steenbergen, 1999). The coordination of arm, neck, and trunk motions establishes the coupling of whole-body movements across the two feeding phases. The other PCs were defined as theorical EMs for the first time. Notably, PCs 3 and 6 indicated shoulder or hip joint fixation in most feeding phases, highlighting the stabilization of proximal body parts. Stability is crucial for the functional performance of the upper extremities and reportedly enhances neutral trunk position (Gillen et al., 2007) and shoulder and trunk fixation (Olczak, Truszczyńska-Baszak, & Mróz, 2022) in individuals with disabilities. Therefore, healthy feeding likely requires stabilizing the upper arm and trunk.

Our PCA revealed that various body segment movements dimensionally reduced the configuration of normal feeding: PC 2 and PC 3 in the reaching phase, PC 2, PC 3, and PC 5 in the spooning phase, PC 3 and PC 5 in the transport phase, and PC 4 in the mouth phase. These upper-body joint motions, which configure each PC in the corresponding feeding phases, have been confirmed in recent research (Doğan et al., 2019; Nakatake et al., 2021). Each EM can be interpreted as follows: PC 2, which involves positioning the hand by changing the two coupled wrist joint motions, represents the approach and manipulation of an object. PC 3 reflects hand transport away from the trunk or closing the mouth via elbow joint motions, representing an upper limb reaching movement. These movements are well-established functions of the upper extremities (Kapandji, Owerko, & Anderson, 2019). Additionally, the elbow joint motion of PC 3 may be adapted for spooning yogurt. Counterintuitively, food intake into the mouth involved neck lateral flexion (PC 4). Wrist flexion/extension (PC 5) enables manipulating and transporting foods, with these aspects warranting consideration.

The results of this study suggest that normal feeding involves various elements related to neck, trunk, and upper extremity movements and postures across different feeding phases. This objective knowledge clarifies our practical experience and supports more effective interventions for patients with eating difficulties, as outlined in the following implications. Practitioners may assess whether the feeding movements of patients are within normal ranges using EMs identified through PCA in this study. Treatment goals and programs focused on specific EMs can be developed to address feeding difficulties. To enhance feeding movements and postures, practitioners might consider implementing targeted positioning strategies, intensive movement training, or the use of adaptive devices, along with providing education for patients and their caregivers or occupational therapy students.

This study has some limitations, including its focus on right-handed individuals using a spoon to eat yogurt, because of the heterogeneity of dominant upper-limb movements between right- and left-handed individuals (Nelson, Berthier, & Konidaris, 2018). These specific conditions may influence the observed EMs and warrant further research to better understand and validate these findings. Individuals exhibiting EMs outside the normal range, such as the minimum or maximum values of PC 1 during the reaching phase, should be noted in clinical assessments. In this study, we employed a portable inertial motion capture system, which offers flexibility in measurement settings. However, alternative methods, such as single-camera markerless capture (Scott et al., 2022) or visual kinematic observation (Bernhardt, Bate, & Matyas, 1998), may offer additional benefits in clinical practice (Demers & Levin, 2017) and should be evaluated for their applicability.

## Conclusions

The PCA of whole-body kinematic data identified several EMs associated with normal feeding, each corresponding to specific functional phases. These findings provide a theorical foundation for defining normal feeding movements and postures. Further research is warranted to validate the application of these findings to clinical practices related to addressing feeding difficulties.

## Data Availability

All relevant data are within the supplementary table 1 and are available online at https://doi.org/10.1371/journal.pone.0259184.s005.

## References

Bernhardt J, Bate PJ, Matyas TA. 1998. Accuracy of observational kinematic assessment of upper-limb movements. Physical Therapy 78:259–270. DOI: 10.1093/ptj/78.3.259.

Bhattacharjya S, Lenker JA, Schraeder R, Ghosh A, Ghosh R, Mandal S. 2021. Comprehensive needs assessment to ensure appropriate rehabilitation training for community-based workers and caregivers in India. American Journal of Occupational Therapy: Official Publication of the American Occupational Therapy Association 75: 7501205130p1–7501205130p10. DOI: 10.5014/ajot.2021.040097.

Bigand F, Prigent E, Berret B, Braffort A. 2021. Decomposing spontaneous sign language into elementary movements: A principal component analysis-based approach. PLOS ONE 16:e0259464. DOI: 10.1371/journal.pone.0259464.

Boop C, Smith J, Kannenberg K. 2017. The practice of occupational therapy in feeding, eating, and swallowing. American Journal of Occupational Therapy: Official Publication of the American Occupational Therapy Association 71(Supplement_2):7112410015p1–7112410015p13. DOI: 10.5014/ajot.2017.716S04.

Burns MK, Patel V, Florescu I, Pochiraju KV, Vinjamuri R. 2017. Low-dimensional synergistic representation of bilateral reaching movements. Frontiers in Bioengineering & Biotechnology 5:2. DOI: 10.3389/fbioe.2017.00002.

Chinju K, Yamamoto Y, Inada E, Iwashita Y, Sato H. 2024. Analysis of head motions during food intake in Japanese adults using a new motion capture system. Archives of Oral Biology 160:105908. DOI: 10.1016/j.archoralbio.2024.105908.

Ciliz O, Tulek Z, Hanagasi H, Bilgic B, Gurvit IH. 2023. Eating difficulties and relationship with nutritional status among patients with dementia. Journal of Nursing Research: JNR 31:e260. DOI: 10.1097/jnr.0000000000000538.

Cipriano-Crespo C, Rodríguez-Hernández M, Cantero-Garlito P, Mariano-Juárez L. 2020. Eating experiences of people with disabilities: A qualitative study in Spain. Healthcare 8:512. DOI: 10.3390/healthcare8040512.

Daffertshofer A, Lamoth CJC, Meijer OG, Beek PJ. 2004. PCA in studying coordination and variability: A tutorial. Clinical Biomechanics 19:415–428. DOI: 10.1016/j.clinbiomech.2004.01.005.

Demers M, Levin MF. 2017. Do activity level outcome measures commonly used in neurological practice assess upper-limb movement quality? Neurorehabilitation & Neural Repair 31:623–637. DOI: 10.1177/1545968317714576.

Doğan M, Koçak M, Onursal Kılınç Ö, Ayvat F, Sütçü G, Ayvat E, Kılınç M, Ünver Ö, Aksu Yildirim S. 2019. Functional range of motion in the upper extremity and trunk joints: Nine functional everyday tasks with inertial sensors. Gait & Posture 70:141–147. DOI: 10.1016/j.gaitpost.2019.02.024.

Gillen G, Boiangiu C, Neuman M, Reinstein R, Schaap Y. 2007. Trunk posture affects upper extremity function of adults. Perceptual & Motor Skills 104:371–380. DOI: 10.2466/pms.104.2.371-380.

Hugh C, Hugh C. 2009. Research Methods and Statistics in Psychology. 5th ed. Routledge. DOI: 10.4324/9780203769669.

Inada E, Saitoh I, Nakakura-Ohshima K, Maruyama T, Iwasaki T, Murakami D, Tanaka M, Hayasaki H, Yamasaki Y. 2012. Association between mouth opening and upper body movement with intake of different-size food pieces during eating. Archives of Oral Biology 57:307–313. DOI: 10.1016/j.archoralbio.2011.08.023.

Jo E-J, Noh D-H, Kam K-Y. 2020. Effects of contextual interference on feeding training in patients with stroke. Human Movement Science 69:102560. DOI: 10.1016/j.humov.2019.102560.

Kaminski TR. 2007. The coupling between upper and lower extremity synergies during whole body reaching. Gait & Posture 26:256–262. DOI: 10.1016/j.gaitpost.2006.09.006.

Kaminski TR, Bock C, Gentile AM. 1995. The coordination between trunk and arm motion during pointing movements. Experimental Brain Research 106:457–466. DOI: 10.1007/BF00231068.

Kapandji IA, Owerko C, Anderson A. 2019. Physiology of the joints, Volume 1 The Upper Limb. 7th ed. Handspring Publisher LTD.

Klinke ME, Wilson ME, Hafsteinsdóttir TB, Jónsdóttir H. 2013. Recognizing new perspectives in eating difficulties following stroke: A concept analysis. Disability & Rehabilitation 35:1491–1500. DOI: 10.3109/09638288.2012.736012.

Koester K, Troeller H, Panter S, Winter E, Patel JJ. 2018. Overview of Intensive Care Unit-related physical and functional impairments and rehabilitation-related devices. Nutrition in Clinical Practice: Official Publication of the American Society for Parenteral & Enteral Nutrition 33:177–184. DOI: 10.1002/ncp.10077.

Kontaxis A, Cutti AG, Johnson GR, Veeger HEJ. 2009. A framework for the definition of standardized protocols for measuring upper-extremity kinematics. Clinical Biomechanics (Bristol, Avon) 24:246–253. DOI: 10.1016/j.clinbiomech.2008.12.009.

Mlinda SJ, Leyna GH, Massawe A. 2018. The effect of a practical nutrition education programme on feeding skills of caregivers of children with cerebral palsy at Muhimbili National Hospital, in Tanzania. Child: Care, Health & Development 44:452–461. DOI: 10.1111/cch.12553.

Nagao T. 2004. Joint Motion Analysis of the Upper Extremity Required for Eating Activities. Bulletin of Health Sciences Kobe 19:13–31.

Nakatake J, Miyazaki S, Arakawa H, Chosa E. 2024. Normal feeding movements expressed by dimensionality reduction of whole-body joint motions using principal component analysis. medRxiv (accessed on November 30, 2024). DOI: 10.1101/2024.09.14.24313686.

Nakatake J, Totoribe K, Arakawa H, Chosa E. 2021. Exploring whole-body kinematics when eating real foods with the dominant hand in healthy adults. PLOS ONE 16:e0259184. DOI: 10.1371/journal.pone.0259184.

Nelson EL, Berthier NE, Konidaris GD. 2018. Handedness and Reach-to-Place Kinematics in Adults: Left-Handers Are Not Reversed Right-Handers. Journal of Motor Behavior 50:381–391. DOI: 10.1080/00222895.2017.1363698.

Olczak A, Truszczyńska-Baszak A, Mróz J. 2022. Change in the results of motor coordination and handgrip strength depending on age and body position-An observational study of stroke patients and healthy volunteers. International Journal of Environmental Research & Public Health 19:4703. DOI: 10.3390/ijerph19084703.

Philipps J, Reinhart C, Rohde A, Virgil K, Moser C. 2012. Feeding and swallowing. Journal of Occupational Therapy, Schools, & Early Intervention 5:90–104. DOI: 10.1080/19411243.2012.701524.

Scott B, Seyres M, Philp F, Chadwick EK, Blana D. 2022. Healthcare applications of single camera markerless motion capture: A scoping review. PeerJ 10:e13517. DOI: 10.7717/peerj.13517.

Shah S, Vanclay F, Cooper B. 1989. Improving the sensitivity of the Barthel index for stroke rehabilitation. Journal of Clinical Epidemiology 42:703–709. DOI: 10.1016/0895-4356(89)90065-6.

Treger I, Aidinof L, Lehrer H, Kalichman L. 2012. Modified constraint-induced movement therapy improved upper limb function in subacute poststroke patients: A small-scale clinical trial. Topics in Stroke Rehabilitation 19:287–293. DOI: 10.1310/tsr1904-287.

Tricon V, Le Pellec-Muller A, Martin N, Mesure S, Azulay J-P, Vernazza-Martin S. 2007. Balance control and adaptation of kinematic synergy in aging adults during forward trunk bending. Neuroscience Letters 415:81–86. DOI: 10.1016/j.neulet.2006.12.046.

Uniform data system for medical rehabilitation. 1990. Guide for Use of the Uniform Data Set for Medical Rehabilitation. Data Management, S., & Center for Functional Assessment, R. State University of New York at Buffalo.

van Andel CJ, Wolterbeek N, Doorenbosch CAM, Veeger DHEJ, Harlaar J. 2008. Complete 3D kinematics of upper extremity functional tasks. Gait & Posture 27:120–127. DOI: 10.1016/j.gaitpost.2007.03.002.

van der Kamp J, Steenbergen B. 1999. The kinematics of eating with a spoon: Bringing the food to the mouth, or the mouth to the food? Experimental Brain Research 129:68–76. DOI: 10.1007/s002210050937.

von Elm E, Altman DG, Eggar M, Pocock SJ, Gφtzsche PC, Vandenbroucke JP. 2007. STROBE initiative. The Strengthening the Reporting of Observational Studies in Epidemiology (STROBE) statement: Guidelines for observational studies. Lancet 370:1453–1457. DOI: 10.1016/s0140-6736(07)61602-X.

Westergren A, Lindholm C, Mattsson A, Ulander K. 2009. Minimal eating observation form: Reliability and validity. Journal of Nutrition, Health & Aging 13:6–12. DOI: 10.1007/s12603-009-0002-4.

